# Genetic variants of *PIEZO1* associate with COVID-19 fatality

**DOI:** 10.1101/2020.06.01.20119651

**Authors:** C.W. Cheng, V. Deivasikamani, M.J. Ludlow, D. De Vecchis, A.C. Kalli, D.J. Beech, P. Sukumar

## Abstract

Fatality from coronavirus disease 19 (COVID-19) is a major problem globally and so identification of its underlying molecular mechanisms would be helpful. The combination of COVID-19 clinical data and genome sequence information is providing a potential route to such mechanisms. Here we took a candidate gene approach to UK Biobank data based on the suggested roles of endothelium and membrane proteins in COVID-19. We focussed on the *PIEZO1* gene, which encodes a non-selective cation channel that mediates endothelial responses to blood flow. The analysis suggests 3 missense *PIEZO1* single nucleotide polymorphisms (SNPs) associated with COVID-19 fatality independently of risk factors. All of them affect amino acids in the proximal N-terminus of PIEZO1, which is an unexplored region of the protein. By using molecular modelling we predict location of all 3 amino acids to a common outward-facing structure of unknown functional significance at the tips of the PIEZO1 propeller blades. Through genome sequence analysis we show that these SNPs vary in prevalence with ethnicity and that the most significant SNP (rs7184427) varies between 65 to 90% even though the reference amino acid is evolutionarily conserved. The data suggest PIEZO1 as a contributor to COVID-19 fatality and factor in ethnic susceptibility.

## INTRODUCTION

Coronavirus disease 19 (COVID-19) is caused by SARS-CoV-2 (severe acute respiratory syndrome coronavirus 2)^1^. In 2020 this virus triggered a global health crisis as it spread rapidly in the absence of a vaccine and with only partially-effective treatments available^2,3^. One way to address such a problem could be to understand how the virus enters host cells because it could usefully inform repurposing of existing therapeutics and enable development of new therapeutics to reduce the dangers of the virus. There is already evidence that surface membrane proteins such as ACE2 and TMPRSS2 are important in SARS-CoV-2 entry^4^ but there is little information on the roles of other membrane proteins, membrane lipid components or membrane structure. Research on other viruses has suggested that such mechanisms affect viral entry^5,6^. Membrane proteins of interest are ion channels^7,8^, which embed in the membrane and allow transmembrane flux of ions such as Ca^2+^, which is a key intracellular signal^9^ and known regulator of coronavirus mechanisms^10-12^.

Investigation of COVID-19 patients has pointed to the importance of endothelial cell infection and endotheliitis^13^. The study included study of the mesenteric vasculature because of circulatory collapse in which there was mesenteric ischaemia requiring surgical resection to remove part of the small intestine^13^. Viral elements were found in endothelial cells and there were accumulations of inflammatory cells alongside endothelial and inflammatory cell death^13^.

It was suggested that therapeutic strategies aimed at stabilising the endothelium may be useful, especially in people who are vulnerable because of pre-existing endothelial dysfunction, such as males, smokers, obese individuals and people with hypertension or diabetes^13^. Similarly, an independent study of lungs from patients who died from COVID-19 found severe endothelial injury and angiogenesis, contrasting with the lungs of patients who died from acute respiratory distress syndrome secondary to influenza^14^. In support of these conclusions, SARS-CoV-2 was found to infect human blood vessel organoids^15^. Therefore, the idea exists that vascular endothelium is particularly vulnerable to SARS-CoV-2 and important in COVID-19 severity^13,14,16,17^.

For these reasons it could be helpful to learn specifically about the membrane mechanisms of endothelial cells that confer SARS-CoV-2 susceptibility and determine downstream consequences of the virus in the vasculature. In 2014, an intriguing ion channel protein called PIEZO1 was reported to be important in endothelium^18,19^. It forms Ca^2+^-permeable non-selective cation channels with extraordinary capability to respond to membrane tension^20^ and shear stress caused by fluid flow along the endothelial membrane surface^18^. Unusually for membrane proteins, PIEZO1 indents the membrane in an inverted dome-like fashion and therefore modifies the overall structure of the membrane^21,22^. The channel shows striking activity in mesenteric endothelium^23^ and there is increasing evidence of its roles in many aspects of endothelial function, such as angiogenesis^18,24^ and pulmonary vascular permeability^25,26^. It also has roles in cardiovascular health and disease more generally^27^, including in the regulation of interleukin-6^28^, which is a key inflammatory mediator of COVID-19^29^. Importance is known in humans because naturally occurring loss-of-function mutations in *PIEZO1* associate with lymphatic endothelial dysfunction^30,31^ and varicose veins^32^, and gain-of-function mutations associate with anaemia and malarial protection due to importance of *PIEZO1* in red blood cells^33-35^. Preliminary genetic analysis has hinted at numerous additional conditions that may be related to mutations in *PIEZO1^27^*.

Therefore, we hypothesised that mutations in *PIEZO1* might relate to COVID-19 and so we tested this hypothesis using data in the UK Biobank. UK Biobank is a health research resource that recruited over 500,000 people aged 40 to 69 years between 2006 and 2010 across the UK^36^. Recently it incorporated COVID-19 infection and fatality data.

## RESULTS

### Demographic analysis

UK Biobank released data on 13502 people tested for COVID-19 across 22 assessment centres as of 3^rd^ August 2020, starting with a first data release on 16^th^ April 2020. Here we carried out a candidate gene association study on these data to identify potential links between COVID-19 and mutations in the *PIEZO1* gene. We elected to focus on 11235 individuals who self-identified as “British” because it presented the largest ethnically similar population. Table 1 shows the demographic properties of the study participants. Of these people, 1294 (11.5%) tested positive. The mean age of all individuals was 70.1 years and 5547 individuals (49%) were men while 5688 (51%) were women. 12% and 11% of men and women tested positive for the virus. The positive cases were further categorised into fatal and non-fatal, with fatal indicating the individuals who were considered to have died due to COVID-19 based on data linking to the National Death Registries Database. Among those who tested positive, 165 (12.8%) were reported to have died due to COVID-19, while 1129 (87.2%) did not die (nonfatal). We evaluated the co-morbidities of these individuals obtained from non-cancer illness code (ID 20002). Of the 11235 individuals, 3861 (34%) self-reported hypertension, 479 (4%) myocardial infarction, 279 (2%) stroke, 800 (7%) diabetes mellitus, 335 (3%) arthritis and 1536 (14%) asthma.

**Table 1.**
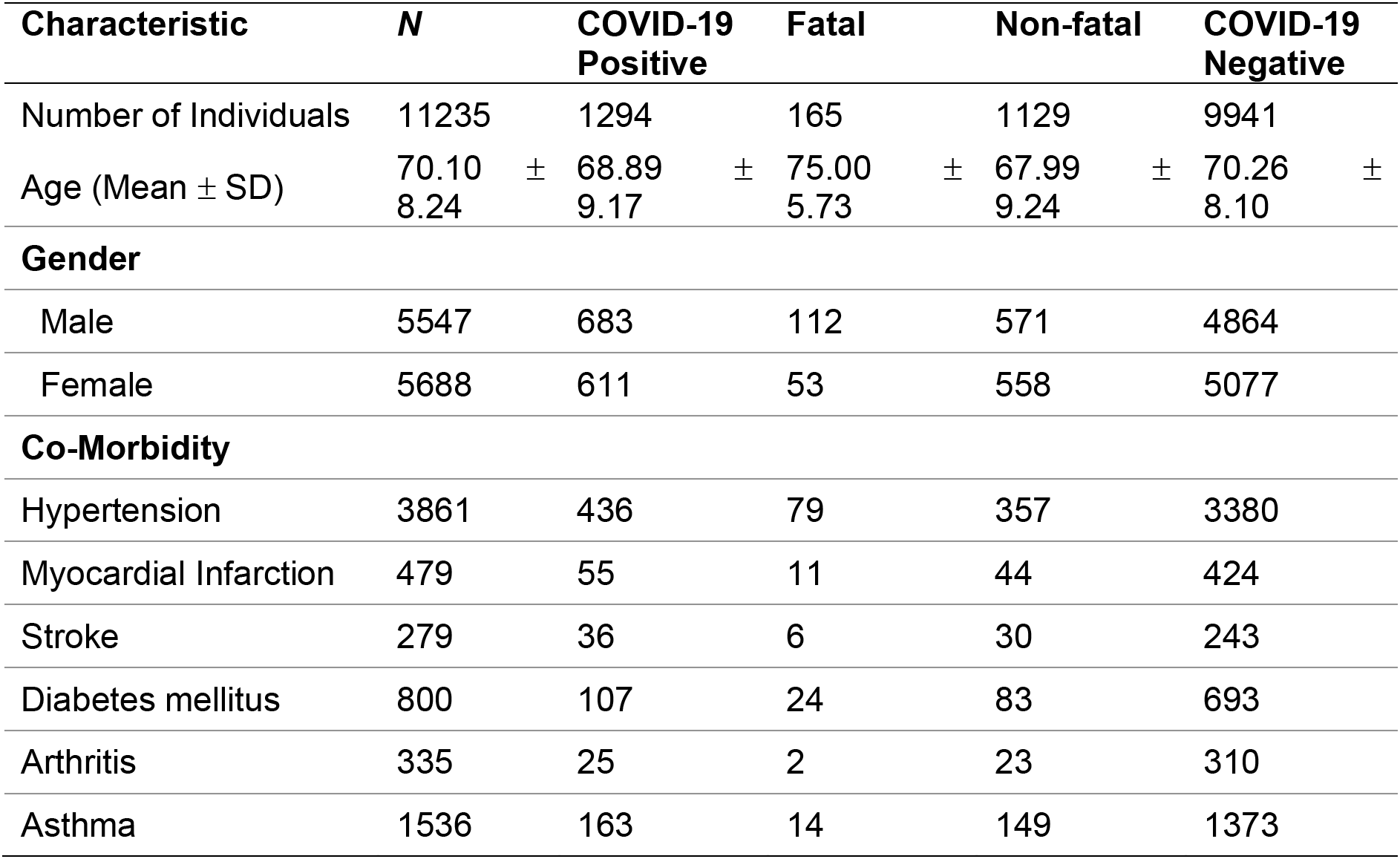
Descriptive statistics and demographic data for COVID-19 in UK Biobank. *N* = Total number of individuals tested for SARS-CoV-2 in UK Biobank; SD = standard deviation. The study focused only on individuals who identified themselves as “British”. The death records were provided by UK Biobank linked to the National Death Registries database. Nonfatal indicates the individuals who were tested positive at least once for SARS-CoV-2 but did not die. Co-morbidities were obtained from self-reported non-cancer illness codes (Field-ID 20002).

### Association analysis strategy

Logistic regression analysis was performed, assuming an additive genetic approach. The first release for 1158 people was classified as the Discovery cohort and the data for all 11235 cases as the Combined cohort. As summarised in Figure 1, the first analysis step (Step 1) was for all individuals who tested positive (“cases”) or negative (“controls”). Then, to identify *PIEZO1* variant linkages to disease severity, analysis was performed as per Steps 2-4: (2) fatal versus negative; (3) non-fatal versus negative; and (4) fatal versus non-fatal. All analyses were adjusted for age, sex, duration of moderate physical activity and the first 10 principal components. Associations with COVID-19 risks were quantified using odds ratios (ORs) derived from logistic regression. Tables 2-6 show the synonymous and missense variants that were detected as statistically significant in at least one of the analyses.

**Figure 1.**
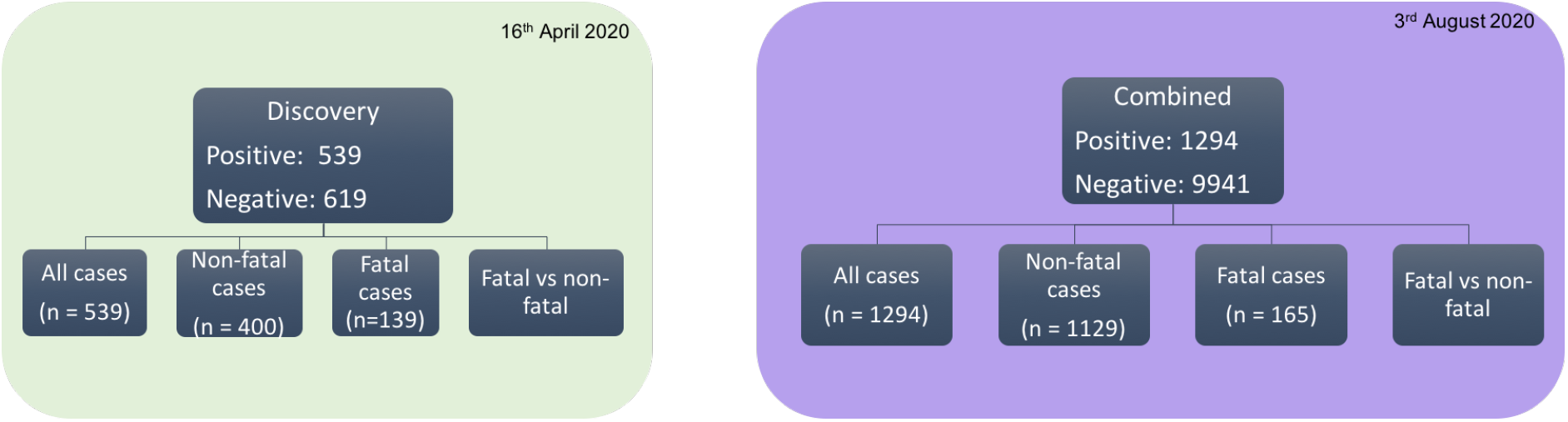
Analysis pipeline. The analyses were restricted to self-reported “British” population. Fatal indicates the death recorded as due to COVID-19. Positive: individuals who tested positive for SARS-CoV-2; Negative: individuals who tested negative for SARS-CoV-2.

**Table 2.** Variation in PIEZO1 amino acid sequence has no effect on susceptibility to infection. Step 1 analysis compared *PIEZO1* in people who tested positive or negative for the virus. SNP, single nucleotide polymorphism; REF = reference allele; ALT = alternate allele; A1 = tested allele; OR = odds ratio; CI = confidence intervals. All analyses were restricted to individuals who self-identified as “British” and adjusted to age, sex, duration of moderate activity, and principal components 1 to 10.

| SNP ID | REF | ALT | A1 | Mutation | Discovery |  | Combined |  |
| --- | --- | --- | --- | --- | --- | --- | --- | --- |
|  |  |  |  |  | *P* | OR<br>(95% CI) | *P* | OR<br>(95% CI) |
| rs8057031 | C | G | C | Synonymous | **0.0048** | 0.7592<br>0.6269 – 0.9193 | 0.5403 | 0.9712<br>0.8845 – 1.0664 |
| rs6500491 | T | C | T | Synonymous | **0.0055** | 0.6344<br>0.4600 – 0.8750 | 0.4436 | 0.9412<br>0.8061 – 1.0090 |
| rs2290902 | A | G | A | Synonymous | **0.0081** | 0.6522<br>0.4753 – 0.8951 | 0.5907 | 0.9588<br>0.8224 – 1.1178 |
| rs1061228 | G | A | A | Synonymous | **0.0114** | 1.4398<br>1.0855 – 1.9098 | 0.5661 | 1.0411<br>0.9072 – 1.1949 |
| rs8043924 | G | C | G | Synonymous | **0.0387** | 0.7299<br>0.5416 – 0.9837 | 0.4364 | 0.9441<br>0.8167 – 1.0913 |
| rs4782430 | A | G | A | Synonymous | **0.0433** | 0.7309<br>0.5394 – 0.9906 | 0.4675 | 0.9471<br>0.8180 – 1.0966 |

**Table 3.**
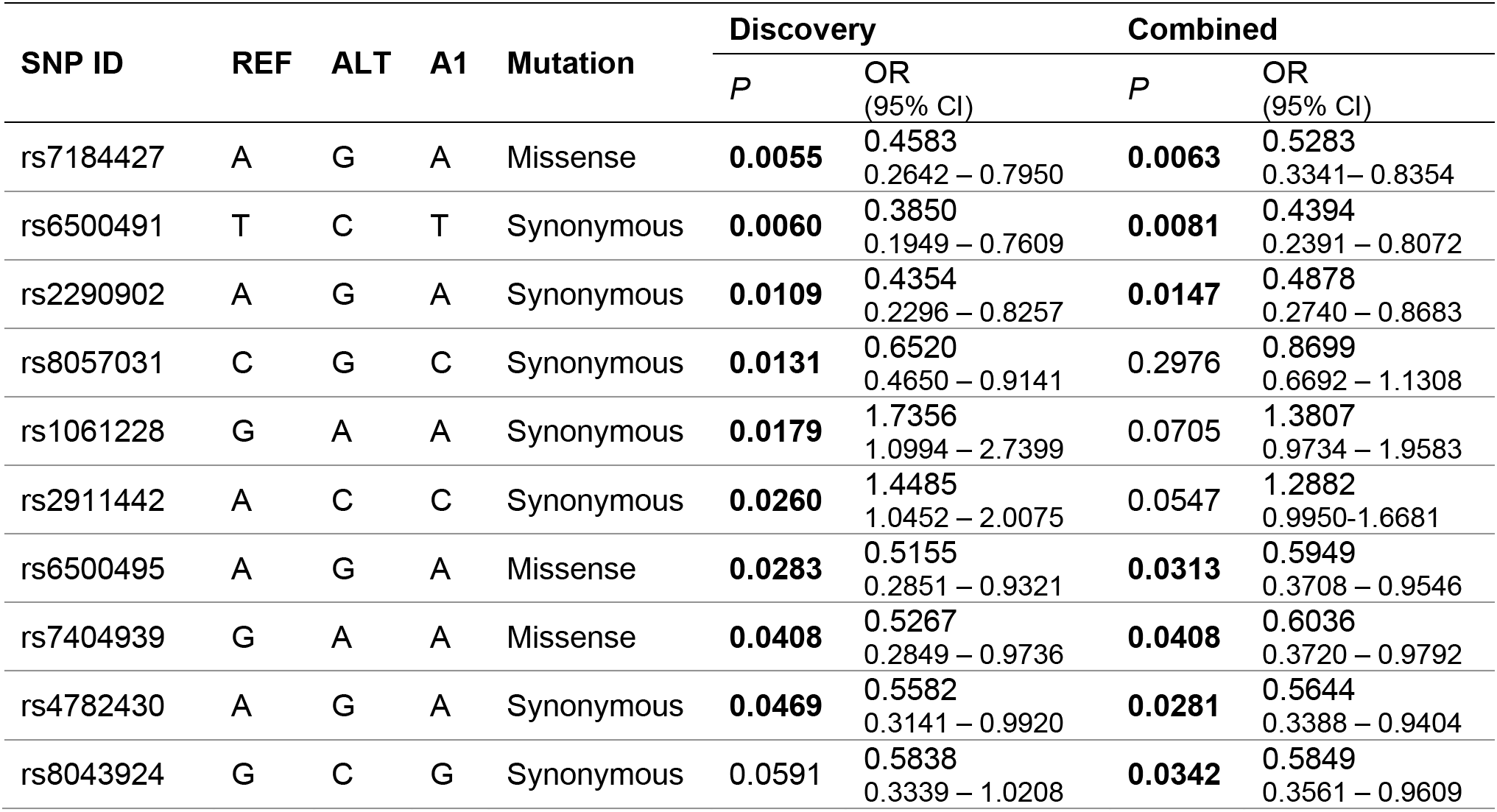
PIEZO1 relationship to COVID-19 fatality: Step 2 association analysis (Fatal versus Negative). SNP, single nucleotide polymorphism; REF = reference allele; ALT = alternate allele; A1 = tested allele; OR = odds ratio; CI = confidence intervals. All analyses were restricted to individuals who self-identified as “British” and adjusted to age, sex, duration of moderate activity, principal component 1 to 10.

**Table 4.**
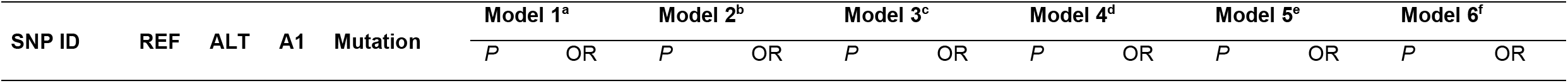
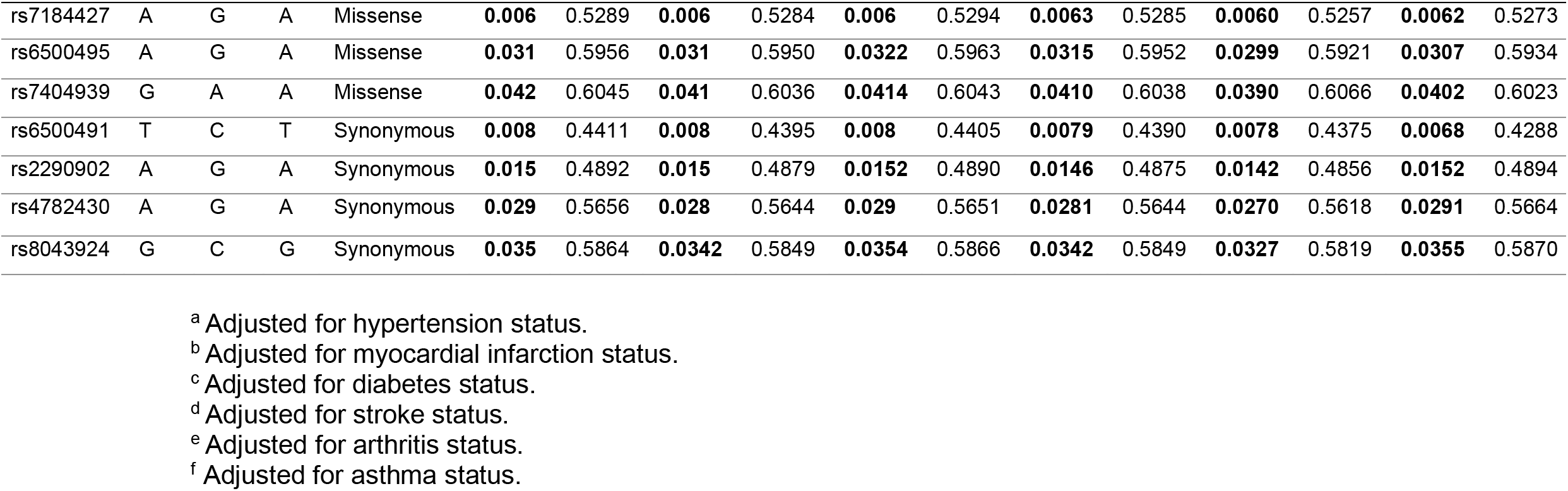
PIEZO1 relationship to COVID-19 fatality: Step 2 association analysis (Fatal versus Negative after adjusting for self-reported illnesses). SNP, single nucleotide polymorphism; REF = reference allele; ALT = alternate allele; A1 = tested allele; OR = odds ratio; all models data were adjusted for age, sex, duration of moderate activity, principal component 1 to 10 in addition to the specific disease condition as given below.

**Table 5.**
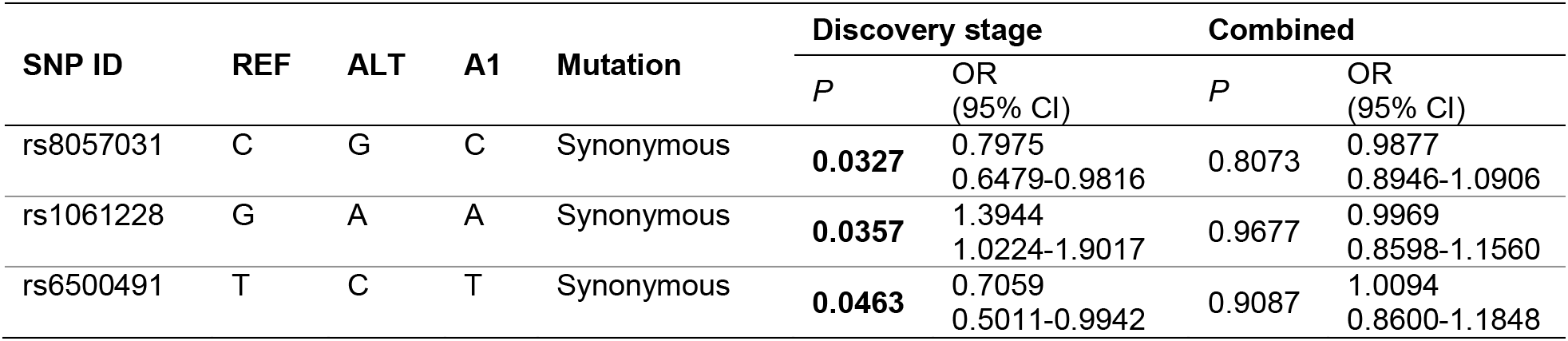
*PIEZO1* has no relationship to non-fatal COVID-19: Step 3 association analysis (Non-fatal versus Negative). SNP, single nucleotide polymorphism; REF = reference allele; ALT = alternate allele; A1 = tested allele; OR = odds ratio; CI = confidence intervals. All analyses were restricted to individuals who self-identified as “British” and adjusted to age, sex, duration of moderate activity, principal component 1 to 10. See also the Supplementary file.

| SNP ID | REF | ALT | A1 | Mutation | Discovery stage |  | Combined |  |
| --- | --- | --- | --- | --- | --- | --- | --- | --- |
|  |  |  |  |  | *P* | OR (95% CI) | *P* | OR (95% CI) |
| rs8057031 | C | G | C | Synonymous | **0.0327** | 0.7975<br>0.6479-0.9816 | 0.8073 | 0.9877<br>0.8946-1.0906 |
| rs1061228 | G | A | A | Synonymous | **0.0357** | 1.3944<br>1.0224-1.9017 | 0.9677 | 0.9969<br>0.8598-1.1560 |
| rs6500491 | T | C | T | Synonymous | **0.0463** | 0.7059<br>0.5011-0.9942 | 0.9087 | 1.0094<br>0.8600-1.1848 |

**Table 6.** PIEZO1 relationship to COVID-19 fatality: Step 4 association analysis (Fatal versus Non-fatal). SNP, single nucleotide polymorphism; REF = reference allele; ALT = alternate allele; A1 = tested allele; OR = odds ratio; CI = confidence intervals. All analyses were restricted to individuals who self-identified as “British” and adjusted to age, sex, duration of moderate activity, principal component 1 to 10.

| SNP ID | REF | ALT | A1 | Mutation | Discovery stage |  | Combined |  |
| --- | --- | --- | --- | --- | --- | --- | --- | --- |
|  |  |  |  |  | P | OR (95% CI) | P | OR (95% CI) |
| rs6500491 | T | C | T | Synonymous | 0.1426 | 0.5792<br>0.2791-1.2019 | **0.0222** | 0.4709<br>0.2469-0.8981 |
| rs2290902 | A | G | A | Synonymous | 0.2127 | 0.6473<br>0.3266-1.2830 | **0.0396** | 0.5266<br>0.2858-0.9700 |
| rs7184427 | A | G | A | Missense | 0.0991 | 0.6303<br>0.3642-1.0909 | **0.0414** | 0.6032<br>0.3711-0.9806 |

### PIEZO1 coding variation has no effect on susceptibility to infection

As indicated, our Step 1 analysis compared people who tested positive for the virus with people who tested negative. In the Discovery cohort (*N* = 1158), 6 synonymous variants were significantly different between the Positive and Negative groups (*P*<0.05) and 26 intronic variants (Table 2 and Supplementary file). But in the Combined cohort, the synonymous variants were not significantly different (*P*>0.05) and only 3 intronic variants remained significant (Supplementary file). The data suggest a possible role for variation in *PIEZO1* as a factor in susceptibility to infection but argue against importance of variation in the PIEZO1 amino acid sequence.

### PIEZO1 variation affects COVID-19 fatality

We next compared data for people who were recorded as having died from COVID-19 (fatal) with data for people who tested negative (Step 2 association analysis: Fatal versus Negative). Analysis was performed on the Discovery cohort (N = 139 Fatal and *N* = 619 Negative) and the Combined cohort (*N* = 165 Fatal and *N* = 9941 Negative). In the Discovery cohort, 67 *PIEZO1* variants were identified as significantly different (*P*<0.05), out of which 3 were missense, 6 synonymous and 58 intronic mutations (Table 3 and Supplementary file). The Combined cohort supported significance for the same 3 missense variants. The most significantly different missense variant in both cohorts was rs7184427, which showed odds ratios (ORs) of 0.4583 and 0.5283 in the Discovery and Combined cohorts respectively. For the synonymous variants, only 3 of the significant differences were for the same residues as the Discovery cohort, and 1 was new (Table 3). The data point to variation in the PIEZO1 amino acid sequences as important in COVID-19 fatality.

To extend this analysis, the Combined cohort data were adjusted to six underlying disorders: hypertension, myocardial infarction, diabetes mellitus, stroke, arthritis and asthma. The purpose of adjusting was to account for covariate effects, thus exploring whether the identified variants reflect true associations between *PIEZO1* and COVID-19 fatality or associations linked to these other diseases. COVID-19 fatality was the response variable while the imputed genotype was the predictor variable used in the PLINK 2.0 logistic regression model. All the models were adjusted to age, sex, duration of moderate physical activity, along with the first ten principal components to control population stratification. Consistently across six models of logistic regression, the results supported relevance of the same 3 missense variants: rs7184427, rs6500495 and rs7404939 (Table 4). In addition, 4 synonymous variants achieved significance (Table 4). The ORs and P-values of each variant did not deviate obviously across the 6 disease models (Table 4). The analysis suggests direct association between variation in PIEZO1 amino acid sequence and COVID-19 fatality.

### *PIEZO1* variation is not associated with non-fatal COVID-19

To investigate if non-fatal infection is linked to *PIEZO1* variants we carried out Step 3 association analysis in which we compared people who tested positive for the virus but did not die (non-fatal cases) with people who tested negative. In the Discovery cohort, consisting of 400 non-fatal cases and 619 people testing negative, 3 synonymous variants were significantly different (Table 5), as were 8 intronic variants (Supplementary file). However, there were no significantly different variants in the Combined cohort (Supplementary file). The data suggest that *PIEZO1* variation lacks relevance to non-fatal COVID-19.

### rs7184427 is consistently associated with fatality

As a further test, we compared fatal and non-fatal cases. In the Discovery cohort, 12 intronic variants were significantly different but no other significant variants were detected (Supplementary file). However, in the Combined cohort there were significant differences for 1 missense and 2 synonymous variants (Table 6) and 10 intronic variants (Supplementary file). The missense variant was the rs7184427 SNP, as identified in the earlier analysis (Tables 3 and 4). The data further suggest importance of PIEZO1 variation in COVID-19 fatality and point particularly to rs7184427.

### rs7184427 affects a highly conserved amino acid residue

The analysis above suggests 3 missense SNPs linked to COVID-19 fatality: rs7184427, rs6500495 and rs7404939; with rs7184427 being particularly strongly implicated. Intriguingly, all of these SNPs affect amino acids in the N-terminus of PIEZO1, about which there is little prior knowledge. Human PIEZO1 comprises 2521 amino acids in total and rs6500495 affects position 83, rs7404939 position 152 and rs7184427 position 250. Therefore, these amino acids lie in the first 10% of the PIEZO1 sequence.

To learn more about this part of the protein, 16 PIEZO1 orthologous protein sequences were compared by constructing a maximum likelihood tree using RAxML (Figure 2A). It shows that: at position 83, isoleucine (I) is unique to humans, whereas in the other species it is threonine (T); at position 152, the residue is highly diverse across species; and at position 250, there is complete conservation of valine (Figure 2B). It should be noted that the analysis was based on reference sequences that do not necessarily reflect the mutations implicated in our UK Biobank analysis: rs6500495 encodes a switch at position 83 from the reference isoleucine to threonine; rs7404939 encodes the reference proline rather than leucine at position 152; and rs7184427 encodes alanine rather than the reference valine at position 250. In summary, the analysis suggests that rs7184427 affects a residue that is highly evolutionarily conserved and therefore likely to have functional importance.

**Figure 2.**
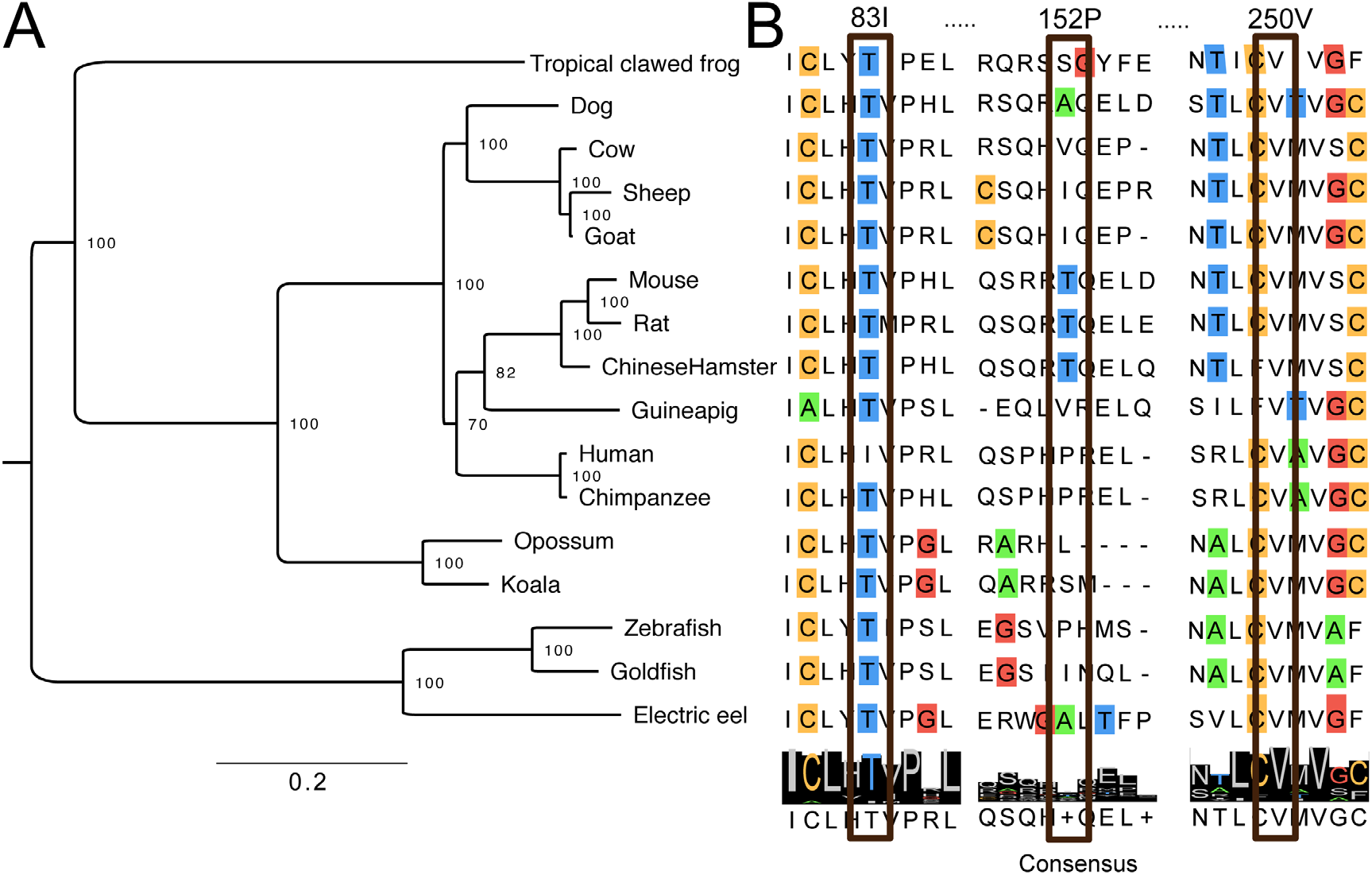
SNP rs7184427 affects a highly conserved amino acid residue. Phylogenetic tree of PIEZO1 sequences across 16 species. (**A**) The phylogenetic tree was inferred from maximum likelihood in RAxML using 100 bootstrap replicates. The tree was midpoint rooted with support values showing on the branch. All nodes achieved >70% bootstrap support. Scale bar represents 0.2 substitutions per site. (**B**) Sequence alignment for amino acids (single letter code) at and near to the residues encoded by rs6500495, rs7404939 and rs7184427. The consensus amino acids are shown at the bottom.

### Prediction of an outward-facing interaction site

Because so little is known about the proximal N-terminus of PIEZO1, we sought insight through molecular modelling of the channel. For these studies we used mouse PIEZO1 because cryo-EM structural data exist for this PIEZO^21^, in which the equivalents of I83, P152 and V250 are T83, T152 and V257 (Figure 3). Unfortunately, the structural data do not include residues 1-576 and thus not residues 83, 152 and 257. However, the similarity and repetitive nature of nearby regions suggested the possibility to predict the entire protein structure, which we did (Figure 3)^22^. Validity of the model was supported by close overlay with the related mouse PIEZO2^22^, for which complete experimentally-derived structural data exist^37^. Viewing our modelled PIEZO1 from the extracellular space shows the expected trimeric arrangement of the 3 PIEZO1s required to form a PIEZO1 ion channel (Figure 3A). The ion pore, which selects for transmembrane flux of cations, is in the centre, and over this lies a large cap, referred to as the C-terminal extracellular domain (CED) (Figure 3A). Spiralling outwards are three long propeller blade-like structures (Figure 3A). T83, T152 and V257 reside at the far-reaches of these blades, distant from the ion pore - each repeated 3 times (Figure 3A). Intriguingly they all lie at the same outer face of the blade tips, ideally placed for interaction with another protein or proteins (Figure 3A). To the best of our knowledge, no such interaction or interactions are actually known and so their existence is speculation. A close-up side view of one distal N-terminal blade shows T83 at the extracellular face of the membrane-spanning structure and T152 and V257 at the cytosolic face (Figure 3B). Overall, the modelling supports the expected distal location of the 3 amino acids affected by missense SNPs and suggests the potential for a common role in an outward-facing interaction site.

**Figure 3.**
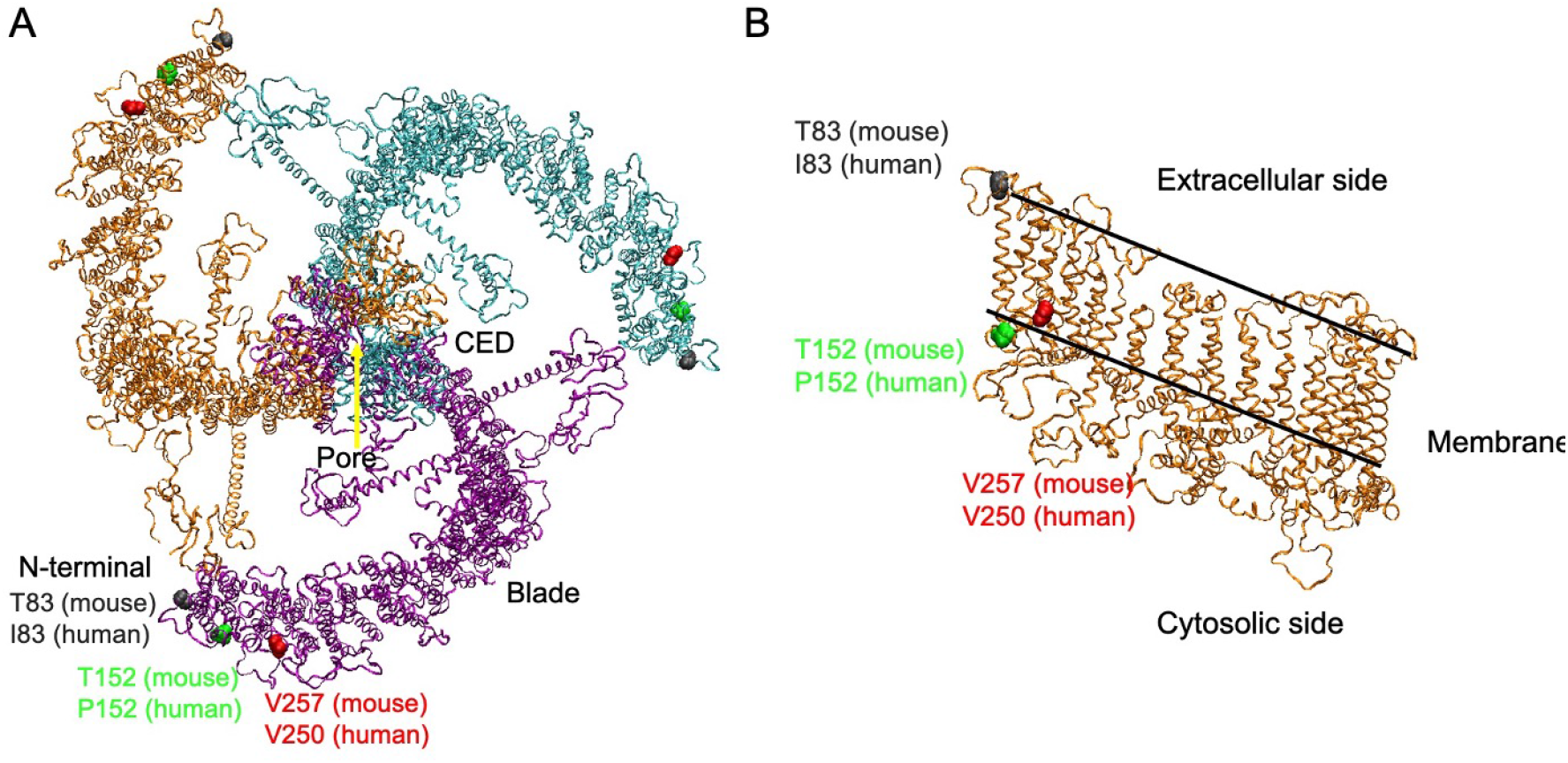
Predicted location of the residue affected by rs6500495, rs7404939 and rs7184427 in the PIEZO1 channel. **(A)** Structural model of the full-length mouse PIEZO1 channel from extracellular side. The three identical PIEZO1 subunits required to make a single PIEZO1 channel are shown in purple, orange and cyan. Isoleucine 83 (Threonine in mouse; black), Proline 152 (Threonine in mouse; green) and Valine 257 (the mouse equivalent of V250; red) are shown. (**B**) Side view showing a close-up of one distal N-terminal region.

### Ethnic variation in prevalence of the missense variants

Suggestions have been made that susceptibility to COVID-19 varies according to ethnicity, partly due to socioeconomic differences but potentially also because of biological variations^38-40^. We therefore investigated whether the pro-disease allele frequency of the 3 significant missense variants (rs7184427, rs6500495 and rs7404939) differs across ethnic groups by analysing gene sequence and ethnicity data in UK Biobank, 1000 Genome and Genome Aggregation Database (gnomAD).

In this analysis it can been seen that the major allele (G allele) frequency of rs7184427 (which is associated with greater COVID-19 fatality and results in V250A mutation) has a generally high frequency of occurrence across all global populations of 82.0-84.4% (Table 7). Therefore, although the reference PIEZO1 sequence indicates valine at this position (Figure 2B), alanine is most common. Analysis by ethnic group shows that the G allele frequency is lower in “South Asian” populations ("Bangladeshi”, “Indian”, “Pakistani”) at 65.0-70.8% (Table 7). In UK Biobank, the frequency is the same for “African” and “British” but higher for “Caribbean”. In GnomAD, where the number of “African” contributors is greater, the G allele frequency is greater than the global average (Table 7). The data suggest that rs7184427 is more common in “African” and “Caribbean” groups where it could potentially result in great fatality, whereas the opposite is the case with “South Asian” ethnicity. Similarly, for rs6500495, the major allele (G allele) frequency is lower in “South Asian” groups and higher in “African” and “Caribbean” groups compared with the global average or “British” group (Table 7). Whereas for rs7404939, the major allele (G allele) frequency is lower in “South Asian” groups and for “African” and “Caribbean” groups compared with the global average or “British” group (Table 7). The data suggest ethnic variation in the frequency of occurrence of the 3 missense PIEZO1 variants.

**Table 7.** Pro-COVID-19 fatality allele frequency for the 3 significant missense variants.

| Population | rs7184427<br>(G allele) | rs6500495<br>(G allele) | rs7404939<br>(G allele) |
| --- | --- | --- | --- |
| **UK Biobank** |  |  |  |
| Global (*n* = 487,410) | 0.844 | 0.8723 | 0.8757 |
| African (*n* = 3,211) | 0.845 | 0.9664 | 0.7312 |
| Bangladeshi (*n* = 223) | 0.705 | 0.8491 | 0.7883 |
| British (*n* = 430,630) | 0.846 | 0.8710 | 0.8806 |
| Caribbean (*n* = 4,303) | 0.879 | 0.9540 | 0.7227 |
| Chinese (*n* = 1,506) | 0.878 | 0.9545 | 0.9555 |
| Indian (*n* = 5,665) | 0.696 | 0.8281 | 0.7768 |
| Pakistani (*n* = 1,750) | 0.708 | 0.8402 | 0.7905 |
| **1000 Genome Database** |  |  |  |
| Global (*n* = 3,500) | 0.820 | 0.909 | 0.822 |
| African (*n* = 1,018) | 0.871 | 0.958 | 0.688 |
| American (*n* = 535) | 0.823 | 0.909 | 0.890 |
| East Asian (*n* = 617) | 0.901 | 0.959 | 0.961 |
| European (*n* = 669) | 0.837 | 0.857 | 0.869 |
| South Asian (*n* = 661) | 0.650 | 0.847 | 0.765 |
| **Genome Aggregation Database (GnomAD)** |  |  |  |
| Global (*n* = 71,702) | 0.829 | 0.888 | 0.838 |
| African (*n* = 21,042) | 0.884 | 0.956 | 0.737 |
| American (*n* = 6,835) | 0.839 | 0.911 | 0.882 |
| Ashkenazi Jewish (*n* = 1,662) | 0.863 | 0.883 | 0.890 |
| East Asian (*n* = 1,567) | 0.880 | 0.956 | 0.948 |
| Finnish (*n* = 5,244) | 0.852 | 0.874 | 0.867 |
| Non-Finnish European (*n* = 32,299) | 0.853 | 0.879 | 0.883 |
| Others (*n* = 1,077) | 0.851 | 0.890 | 0.859 |
| South Asian (*n* = 1,526) | 0.693 | 0.836 | 0.777 |

### Synonymous variants also differ in frequency with ethnicity

Similar analysis was performed for synonymous mutations associate with COVID-19 fatality in at least one step of the association analysis (Figure 4). Focussing only on those that were significant in the Combined cohort after adjusting for self-reported illnesses: rs4782430 is also more frequent in African and Caribbean groups and less frequent in South Asian groups; rs8043924 is also less frequent in South Asian groups; but rs2290902 and rs8043924 are less frequent in African and Caribbean groups; and rs6500491 shows no differences (Figure 4). The data suggest that the synonymous variants also vary in frequency with ethnicity but that the pattern is not consistent for all mutations.

**Figure 4.**
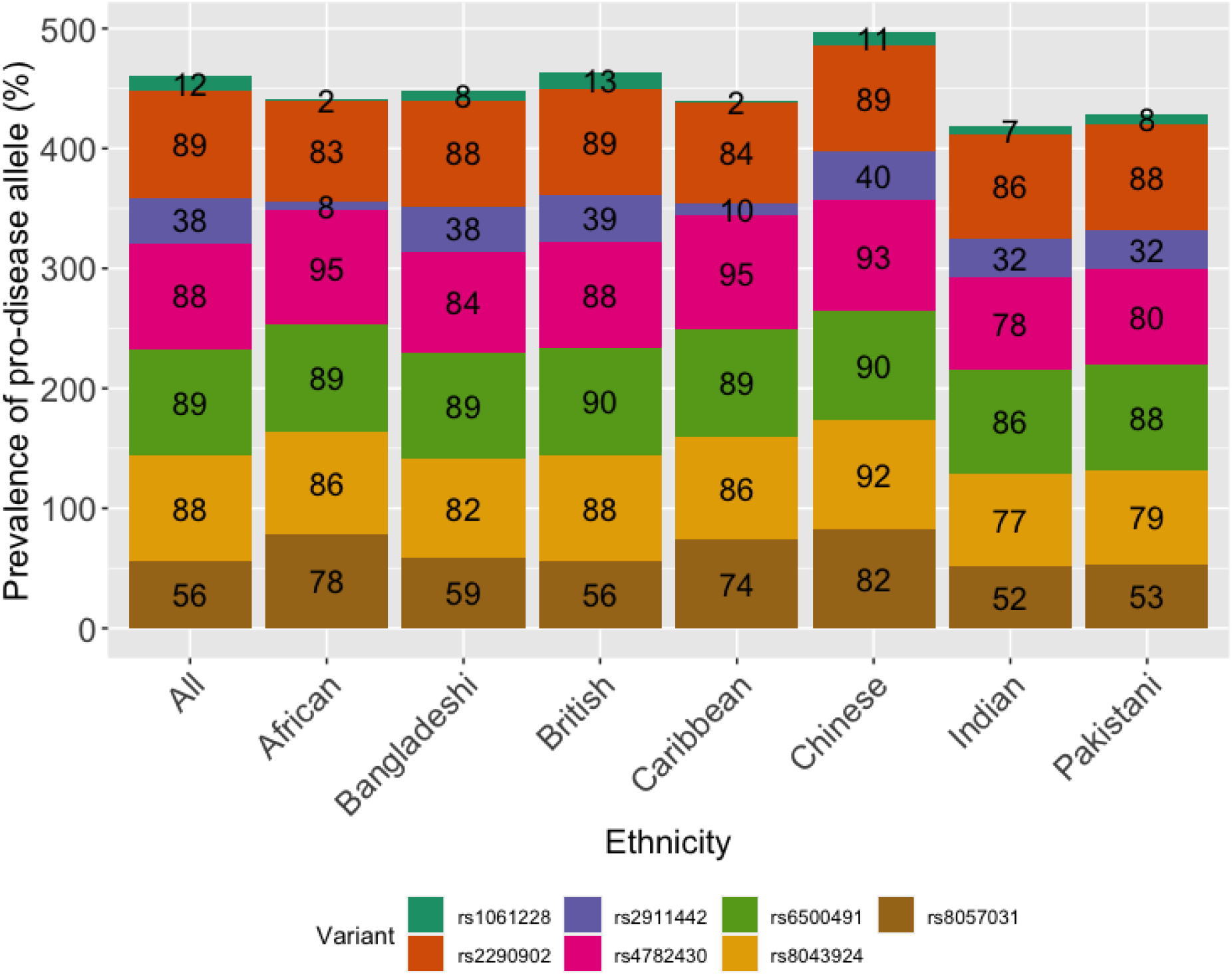
Pro-COVID-19 fatality allele frequency of synonymous mutations across different populations in UK Biobank (expressed as percentages).

## DISCUSSION

The study suggests that genetic variation in *PIEZO1* is a factor in COVID-19 fatality. One particular variant stands out: rs7184427. This SNP is a missense mutation that causes a change in amino acid in the PIEZO1 N-terminus. According to phylogenetic analysis of reference sequences, the amino acid at this position is a highly evolutionarily conserved valine. This suggests functional importance, although the nature of this importance is currently unclear. Moreover, although valine is in the reference sequence, rs7184427 causes a switch to alanine. This switch, which associates with the greatest COVID-19 fatality, is the most common variant in humans globally. The frequency varies with ethnicity and may be a factor in the different susceptibilities of different ethnic groups to lethal effects of COVID-19.

Geographic and socio-economic factors account for at least half of the difference in risk of COVID-19 fatality between people of Black and White ethnicity in the UK, but these factors do not explain all of the difference^40^. Greater genetic susceptibility of Black people could be a factor and our analysis lends some support to this hypothesis because rs7184427 appears to be more common in people of African or Caribbean ethnicity. It is notable, however, that rs7184427 is less common in people of South Asian ethnicity even though these people suffer great COVID-19 fatality in the UK^40^.

We do not yet know if alanine in place of valine at position 250 in PIEZO1 causes less, more or no change in the properties of the PIEZO1 channels or whether the altered DNA sequence affects *PIEZO1* expression. The valine amino acid is similar to the alanine amino acid but this chemical difference can be crucial in α-helical integrity^41^.

While we have insight into how some parts of the PIEZO1 molecular machine operate^21,42,43^, the particular region of PIEZO1 incorporating V or A at position 250 is uncharted territory^22^. Based on our molecular modelling we suggest that V250A forms part of an outward-facing domain at the tip of the propeller blades, along with the other residues affected by SNPs (I83 and P152) (Figure 3A). Such a domain seems ideally placed to interact with nearby proteins but could also be important in refining mechanical sensitivity and how the channel interacts with lipids^22^. The molecular and functional studies required to test these ideas will take time to complete. Moreover, our studies have shown the importance of understanding PIEZO1 in its native membrane environment because the native endothelial and red blood cell environments profoundly alter the gating characteristics of PIEZO1^18,27,44,45^. We do not yet know whether endothelial cell dysfunction affects the properties of PIEZO1, but any changes could be relevant to COVID-19 and its progression to fatality.

There is a public database entry indicating a link of V250A to hereditary lymphedema in one patient^46^ and we know that generalised lymphatic dysplasia can be caused by loss of PIEZO1 expression^30,31^. On this basis we might speculate that V250A reduces PIEZO1 function and that this is the reason for increased susceptibility to COVID-19 fatality. If this is the case, PIEZO1 agonists may be a route to rescuing normal channel activity and protecting against fatality. This may be a safe approach because we know that PIEZO1 gain-of-function is tolerated in people and even confers protective advantage in large populations where malaria is endemic^33-35^. A screen of 3.25 million small-molecules identified a PIEZO1 agonist called Yoda1^47^. It is increasingly used successfully as a tool compound in experimental studies of PIEZO1^27^ and there is on-going work to elucidate the chemical structure-activity requirements and improve the physicochemical properties while retaining efficacy; some of these studies have been published^48^. To the best of our knowledge there is currently no PIEZO1 agonist that would be suitable for administration to people but the available data suggest that the principle of chemical enhancement of PIEZO1 is possible and thus that a therapeutic drug targeted to PIEZO1 is a realistic consideration.

In addition to a possible link to lymphedema, our study raises the question of whether COVID-19 is linked to varicose veins. A genome-wide association study identified mutations in *PIEZO1* as determinants of varicose veins^32^ and a recent study specifically linked V250A to varicose veins^49^. These observations further encourage the idea of an important relationship between vascular integrity and COVID-19^13,14,16,17^.

PIEZO1 may act by impacting membrane structure in such a way as to affect viral entry, but we should also consider the property of PIEZO1 to confer Ca^2+^ permeability on the membrane because transmissibility of both SARS-CoV and MERS-CoV is Ca^2+^-dependent and both are closely related to SARS-CoV-2^10,11,50^. E protein of SARS-Co-V forms Ca^2+^ permeable ion pores on ER/Golgi membrane and its activity can drive the inflammation and severe respiratory distress syndrome by enhancing IL-1β production^51^. One of the accessory proteins of SARS-CoV undergoes conformational changes upon binding to Ca^2+^ and significantly contributes to disease progression^12,52^.

Synonymous mutations do not change the amino acid sequence but they could still, in theory, have effects by altering expression of the *PIEZO1* gene, thereby affecting the total amount of PIEZO1 available for functional consequence. We note that one of the synonymous mutations we identified (rs2290902) has also been associated with HIV-1 infection^53^ but we are not aware of further investigation of this matter. It is nevertheless consistent with our general idea that PIEZO1 is relevant to viral disease.

In addition to its endothelial and red blood cell functions, PIEZO1 is expressed in other cell types such as epithelial cells and implicated in other aspects of mammalian biology^27,43^. It may be particularly important to note in this context the recent persuasive evidence for a role of PIEZO1 in pulmonary inflammation, bacterial infection and fibrotic auto-inflammation^54^. In this study it was concluded that stimulation of PIEZO1 in immune cells by cyclical mechanical force is essential for innate immunity. Therefore, we speculate that any loss of PIEZO1 function would increase susceptibility to lung infection and perhaps other infections and that PIEZO1 agonism might be beneficial. Cyclical activation of PIEZO1 might also be important in the protective benefits of physical exercise where PIEZO1 has been suggested to have an important role in blood pressure elevation of exercise that increases performance^23^. PIEZO1 might be relevant to the speculation about a connection between malarial protection and COVID-19^55,56^ because heterozygosity for PIEZO1 gain-of-function mutation has been suggested to be a major factor mediating protection from malaria^35^.

We recognise that a potential limitation of our study is the relatively small size of the COVID-19 data set on which we could base the analysis. Analysis of additional data as they become available will be important for deeper understanding and exploration of *PIEZO1* in all groups and specifically in non-White ethnic groups.

In an initial preprint released on 3 June 2020 based on UK Biobank data available on 16 April 2020 we suggested association of rs7184427 with SARS-CoV-2 infectivity (doi: https://doi.org/10.1101/2020.06.01.20119651; http://dx.doi.org/10.2139/ssrn.3618312). Subsequent to the 16 April release, changes were made to the underlying data by UK Biobank, we presume to improve accuracy. These changes caused the P-value for rs7184427 to become slightly greater than the threshold value for significance in regard to infection rates. Therefore, as reported in this article, we no longer suggest significant association of this variant with infectivity. It is intriguing that this same variant is the one that is most significantly associated with COVID-19 fatality. It may be that larger data sets will again suggest a link to infectivity and that rs7184427 has a role in susceptibility to infection as well as fatality.

In summary, our study suggests genetic association that is relevant to COVID-19 fatality and ethnic variation in susceptibility to fatality and a previously unrecognised relationship between COVID-19 and *PIEZO1*, a gene that encodes an important mechanically-activated ion channel of the endothelium and other cellular structures that are relevant to this disease.

## METHODS

### Demographic information

The UK Biobank resource recruited over 500,000 people aged 40 to 69 years between 2006 and 2010 across the UK^36^. Participants completed a detailed clinical, demographic and lifestyle questionnaire, underwent clinical measures, provided biological samples (blood, urine and saliva) for future analysis and agreed to have their health records accessed. In July 2017, the genetic information from 501,708 samples was released to UK Biobank research collaborators. The UK Biobank has now released the data of individuals tested for COVID-19 in different updates. The first release of the COVID-19 dataset served as the discovery cohort in the present study while the subsequent releases were combined with the discovery cohort to form a combined cohort to study the association between COVID-19 and *PIEZO1*.

*Discovery cohort:* On 16^th^ April 2020, the UK Biobank released the data of 1474 individuals tested for COVID-19 across its 22 assessment centres. Of those, 1158 were British.

*Combined cohort:* This cohort included all individuals from the five different releases by UK Biobank *(N* = 13502). Of those, 11235 individuals were British.

Baseline assessments of all individuals tested for COVID-19 were recorded when they attended one of the 22 research centres located across the United Kingdom. Co-morbidity details of these subjects was taken from the self-reported non-cancer illness codes in the UK Biobank (Data-field 20002), which includes self-reported hypertension, myocardial infarction, stroke, diabetes mellitus, arthritis and asthma. Arthritis status consists of self-reported rheumatoid arthritis and osteoarthritis.

### Statistical analysis

Imputed genotypes of chromosome 16 for all 1158 (discovery cohort) and 11235 (combined cohort) individuals who self-reported as “British” were used for the association study. Imputation was conducted by UK Biobank using IMPUTE2^57^. As this is a candidate gene association study, we focused on the *PIEZO1* gene sequence. Variants with imputation quality (Info) score above 0.4 were retained in the analysis. The variants were further filtered based on minor allele frequency (MAF) >5%, missingness >10%, Hardy-Weinberg equilibrium (HWE) >1×10^-6^ and sample missingness >10%. Of the total number of 197,730 variants spanning chromosome 16, following the filtering parameters above, only 196,314 variants were eligible for the downstream analysis. We analysed 334 variants spanning the coding region of *PIEZO1* and the statistical significance was set at P<0.05 (0.05/1 gene). To reduce the risk of confounding factors due to differences in ancestral background in the UK Biobank, the analyses were restricted to individuals who self-reported as “British” comprising 1158 individuals in discovery cohort and 11235 individuals in combined cohort.

*Step 1 of association analysis*. The first step was conducted on the association between COVID-19 and *PIEZO1* in all positive and negative individuals in both the cohorts. The association analysis was performed using logistic regression implemented in PLINK v2.0 (whole genome data analysis toolset) ^58^. The COVID-19 status was used as a categorical variable in the logistic regression analysis. Individuals tested positive at least once for COVID-19 were considered as cases while the negatives were considered as controls. In the discovery cohort, 539 cases were compared against 619 controls. In the combined cohort, all positive cases comprised of 1294 individuals against 9941 controls. To account for population structure, the first ten principal components were calculated for the cohorts using “--pca” implemented in PLINK 2.0 and used as covariates in the logistic regression. The duration of moderate activity was obtained from the UK Biobank (Field-ID 894) and adjusted as a covariate since *PIEZO1* acts as an exercise sensor, mediating optimised redistribution of blood flow to sustain activity^23^. The activity level was determined using a questionnaire on duration of exercise/activity each day. The logistic regression analyses were also adjusted to sex and age. The workflow of the analysis pipeline is presented in Figure 1.

*Step 2 of association analysis*. To determine whether *PIEZO1* variants were linked with COVID-19 fatality, step 2 of the analysis focused on the deaths due to COVID-19. The death register data were provided by UK Biobank through the National Death Registries database. The cause of death due to COVID-19 was labelled as “U071” in the data-field 40001. In the analysis, only the deaths caused by COVID-19 were categorised as fatal cases while the negatives were classified as controls in the logistic regression analysis. There were 139 fatal cases versus 619 negatives in the discovery cohort and 165 fatal cases versus 9941 negatives in the combined cohort. For the combined cohort, we additionally adjusted the analysis to six self-reported non-cancer illnesses (Field-ID 20002) namely hypertension, myocardial infarction, stroke, diabetes mellitus, arthritis and asthma status. Age, sex, duration of moderate activity and first 10 principal components were used as covariates.

*Step 3 of association analysis*. For step 3 of the analysis, individuals who were tested positive for COVID-19 but not recorded dead were defined as cases. Logistic regression under additive model was used to test the association between non-fatal cases and negatives. In the discovery cohort, there were 400 non-fatal cases against 619 negatives. 1129 non-fatal and 9941 negatives were found in combined cohort. Similarly, the analyses were adjusted to age, sex, duration of moderate activity and first 10 principal components.

*Step 4 of association analysis*. We aimed to identify the *PIEZO1* variants associated with disease severity in COVID-19. The final step of the analysis focused on fatal and nonfatal cases of COVID-19. Using logistic regression under additive model, the cases were defined as deaths due to COVID-19 while the controls were defined as positive for COVID-19 but not dead. There were 139 fatal versus 400 non-fatal in discovery cohort and 165 fatal versus 1129 non-fatal in combined cohort. We controlled for age, sex, duration of moderate activity and first 10 principal components in all the analyses. The analysis pipeline is summarised in Figure 1. In four steps of the analyses, ANNOVAR^59^ was used to annotate the variants to predict missense variants, frameshift mutations and intronic variants.

### Phylogenetic analysis

Protein sequences encoded by the *PIEZO1* gene were obtained from the Ensembl database ^60^. Human *PIEZO1* protein (ENSP00000301015) was used as a reference to retrieve orthologous sequences in other species: chimpanzee (ENSPTRP00000014430), rat (ENSRNOP00000069995), mouse (ENSMUSP00000089777), koala (ENSPCIP00000009017), Opossum (ENSMODP00000052005), goldfish (ENSCARP00000072858), guinea pig (ENSCPOP00000030993), Chinese hamster (SCGRP00001016097), sheep (ENSOARP00000014314), Goat (ENSCHIP00000024918), dog (ENSCAFP00000029440), cow (ENSBTAP00000027897), zebrafish (ENSDARP00000101429), tropical clawed frog (ENSXETP00000090443) and electric eel (ENSEEEP00000041564).

The amino acid sequences of PIEZO1 in these 16 species were aligned using CLUSTAL OMEGA^61^. The resulting alignments were manually curated and trimmed using alignment editor in MEGA 7^62^. Next, the amino acid sequences were subjected to phylogenetic analysis using RAxML^63^ based on maximum likelihood algorithm. The phylogenetic tree was generated with 100 rapid bootstrap replicates. The tree was visualised in Fig Tree version 1.4.3^64^ and the bootstrap support values labelled on the nodes.

### Molecular modelling of PIEZO1

This methodology has been reported previously^22^.

## Data Availability

The data analysed are available to the registered users through UK Biobank

https://www.ukbiobank.ac.uk/

## DATA AVAILABILITY

The data analysed are available to the registered users through UK Biobank.

## SUPPLEMENTAL INFORMATION (SI)

Comprehensive data are provided in the Supplementary file.

## ACKNOWLEDGEMENTS

This research has been conducted using the UK Biobank Resource (Project ID: 42651). The work was supported by a Wellcome Trust Investigator Award to DJB, a BHF Daphne Jackson Fellowship to VD and Leeds Cardiovascular Endowment funds for support of CWC. The study involved use of ARC4, which is part of the High Performance Computing Facility at the University of Leeds. We thank Richard Cubbon for helpful feedback.

## AUTHOR CONTRIBUTIONS

CWC, VD and MJL performed DNA sequence analysis. DDV and ACK performed molecular modelling. CWC, DJB and PS conceived the study. CWC, DJB and PS co-wrote the article with input from all authors. DJB and PS generated research funds and coordinated the project.

